# COVID-19 in rheumatic diseases: A random cross-sectional telephonic survey

**DOI:** 10.1101/2020.08.09.20170985

**Authors:** Rudra Prosad Goswami, Uma Kumar, Maumita Kanjilal, Debaditya Ray, Pallavi Vij, Dheeraj K Mittal, Laxman Meena, Sandeep Nagar, Danveer Bhadu

**Affiliations:** Assistant Professor, Department of Rheumatology, All India Institute of Medical Sciences, New Delhi, India; Professor and Head, Department of Rheumatology, All India Institute of Medical Sciences, New Delhi, India; Research Officer, Department of Rheumatology, All India Institute of Medical Sciences, New Delhi, India; Senior Resident, Department of Rheumatology, All India Institute of Medical Sciences, New Delhi, India

**Author notes:** **Corresponding author** Uma Kumar, Professor and Head, Department of Rheumatology, Room No. 4076, Fourth Floor Teaching Block, All India Institute of Medical Sciences, Ansari Nagar, New Delhi-110029, India.

**Keywords:** Ankylosing Spondylitis, COVID-19, Incidence, SARS-CoV-2, Rheumatoid arthritis, Rheumatology, Systemic lupus erythematosus

## Abstract

**Objective:** To describe the incidence, clinical course, and predictive factors of coronavirus 2019 (COVID-19) infection in a cohort of rheumatological patients residing in New Delhi (National Capital Region), India.

**Methods:** We performed a cross-sectional, random telephonic survey from 20^th^ April to _20_^th^ July 2020 on patients with rheumatic diseases. Patients were interviewed with a predesigned questionnaire. The incidence of COVID-19 in the general population was obtained from open access government data repository. Report of reverse transcriptase polymerase chain reaction report was taken as confirmatory of COVID-19 infection.

**Results:** Among the 900 contacted patients 840 responded (713 with rheumatoid arthritis (RA), 100 with systemic lupus erythematosus (SLE), 20 with spondylarthritis (SpA) and 7 with others; mean age 45 ±13 years, mean duration 11.3 ± 6.3 years; 86% female). Among them 29 reported flu-like symptoms and four RA patients had confirmed COVID-19 infection. All of them were hospitalized with uneventful recovery. Rheumatological drugs were discontinued during the infectious episode. Disease modifying agents and biologics were equally received by those with or without COVID-19. The incidence of COVID-19 was similar to general Delhi population (0.476% vs 0.519% respectively, p=0.86). Two patients had relapse of rheumatic disease after recovery. After recovery from COVID-19 or Flu-like illness, eight patients (27.6%, 95% confidence interval 14.7-45.7) reported disease flare.

**Conclusion:** Patients with rheumatic diseases in India have similar incidence of COVID-19 infection compared to the community. Relapse of underlying rheumatic disease after recovery is not uncommon and continuation of glucocorticoid through the infection should be considered.

## Introduction

The current severe acute respiratory syndrome coronavirus 2 (SARS-CoV-2) pandemic which was declared a public health emergency of international concern on January 2020 has posed a special challenge to rheumatologists [1]. Patients with rheumatic are expected to be at an increased risk of severe infection because of inherent immune dysregulation, associated co-morbidities and, use of immunosuppressive drugs. It is well known that viral infections commonly occur among these patients especially respiratory viral infections [2]. Syndrome of cytokine storm and immune mediated systemic thrombosis have emerged as contributors to the disease severity in COrona VIrus Disease (COVID19) [**3**]. While both conventional (csDMARDs) and targeted disease-modifying antirheumatic drugs (tsDMARDs) increase risk of various infections, some of them are currently being tested as potential therapeutic agent for COVID-19 [4], such as reports of in-vitro viral inhibition with hydroxychloroquine (HCQ) [5] and potential role of tocilizumab in the virus-related cytokine storm [3]. Also, to contain the epidemic there were widespread “lockdowns” in India, resulting in difficulty in getting access to health care facilities which lead to the initiation of “teleconsultation” services. However, an accurate representation of status of patients with rheumatic diseases, during these difficult times of pandemic is definitely lacking but data on epidemiology of SARS-CoV-2 among these patients is scarce but emerging slowly building up confidence amongst rheumatologists. To address this, we conducted cross-sectional, random telephonic survey from 20^th^ April to 20^th^ July 2020 to assess the incidence, and the predictive factors of SARS-CoV-2 infection in a cohort of rheumatological patients residing in Delhi, one of the worst hit regions of India.

### Patients and methods

The present study was a cross-sectional, random telephonic survey conducted from 20^th^ April to 20^th^ July 2020 on patients with rheumatological disorders, under long-term follow-up at the Department of Rheumatology, All India Institute of Medical Sciences, New Delhi, India, and residing in Delhi-NCR (hot spot for COVID19), India. Telephonic method was chosen because of ongoing lockdown and consequent travel restrictions limiting physical access to patients. The study protocol was reviewed and approved by the institutional review board (no. IEC-280/17.4.2020).

Inclusion criteria were: adult patients (>18 years of age) with a definite rheumatic disease diagnosed by a physician and under regular follow up at our Department for more than 1 year. Patients were telephonically contacted and a verbal consent was obtained. Patient data were extracted based on a predesigned proforma which included demographic details, disease duration, current disease status, medication details (details of csDMARDs, tsDMARDs and monoclonal antibodies); travel history (within and outside India); symptoms suggestive of COVID-19 (flu-like symptoms and others with leading questions for anosmia, dysgeusia, red eyes, skin rashes, shortness of breath and dry cough), status of COVID-19 test (whether a nasopharyngeal swab was done or not); history of contact with a COVID 19 positive case and need for hospital admission and course of hospital treatment; and status of rheumatic disease during the period of study. In certain cases, especially in patients with reported increased disease activity, or those who were COVID19 positive, were contacted multiple times throughout the period to ensure the correct clinical course of both COVID-19 and the underlying rheumatic disease.

### Statistical methods

Descriptive data was displayed as mean ± standard deviation and categorical data as percentage (fraction). Comparison of means was done with Mann Whitney U test and comparison of proportions with Fisher’s exact test or Chi Squared test, as appropriate. Incidence of COVID-19 was expressed as percentage with 95% confidence interval (CI). For comparison of incidences across different diagnostic classes, to correct for family wise error rate, correction of p-value divided by number of pre-defined comparisons was taken as cut-off. All statistical calculations were done using Microsoft Excel Spreadsheets and SPSS ver 20 (IBM Corp.).

## Results

### Description of the cohort

Overall, 900 patients were contacted and 840 patients responded. The mean age was 45 ± 13 years with 54% females (n=456). Detailed demographic and clinical details of the cohort is given in Table 1. Majority of the patients had rheumatoid arthritis (RA) (85%, 713/840) followed by systemic lupus erythematosus (SLE) (12%, 100/840) and others had the following diagnoses: ankylosing spondylitis (AS) (n=14), psoriatic arthritis (PsA) (n=6), scleroderma (SSc) (n=3), Sjogren syndrome (SjS) (n=3) and dermatomyositis (n=1). Mean duration of disease was 11.3 ± 6.3 years. Overall, 78% (656/840) were on at least one of prednisolone, csDMARDs, tsDMARDs or on monoclonal antibodies and the rest 184 (22%) patients were only on hydroxychloroquine. Mean doses of the drugs were as follows: prednisolone 5.1 ± 1 mg/day; hydroxychloroquine 321 ± 73 mg/day; methotrexate 18.8 ± 4.3 mg/week; leflunomide 17.4 ± 4 mg/day; sulfasalazine 1.8 ± 0.6 gm/day; mycophenolate mofetil 2000 mg/day and azathioprine 100 mg/day. Approximately one third patients (36%, 302/840) had at least one comorbidity. Fifty-two patients (6%) had ≥2 comorbidities. Sixteen patients reported stoppage of medications because of the fear of contracting the infection or difficulty in procuring drug due to lockdown in India: hydroxychloroquine in nine, methotrexate in four, leflunomide in two and prednisolone, infliximab and tocilizumab in one each. Eleven percent (11/100) of SLE patients reported increased disease activity and 17% (121/713) patient with RA reported increased joint symptoms. Among the 11 lupus disease flares, one was renal, five haematological, two musculoskeletal and three were cutaneous.

**Table 1.** Demographic, clinical and drug related data of the entire cohort, definite COVID-19 and suspected COVID-19 patients

| Parameters |  | Total<br>n=840 | Definite COVID-19 patients<br>n=4 | Suspected COVID-19 patients<br>n=25 |
| --- | --- | --- | --- | --- |
| Age in years | | 45 ± 13 | 40 ± 12.1 | 49 ± 12 |
| Duration of disease in years | | 11.3 ± 6.3 | 8.5 ± 2.7 | 14.4 ± 7.8 |
| Female gender |  | 54 (456) | 75 (3) | 84 (21) |
| History of travel within India |  | 1.5 (13) | 0 | 0 |
| History of travel outside India |  | 0 | 0 | 0 |
| Diagnosis | RA | 85 (713) | 66.6 (4) | 84 (21) |
|  | SLE | 12 (100) | 0 | 12 (3) |
|  | AS | 1.6 (14) | 0 | 4 (1) |
|  | PsA | 0.7 (6) | 0 | 0 |
|  | SSc | 0.3 (3) | 0 | 0 |
|  | SjS | 0.3 (3) | 0 | 0 |
|  | Dermatomyositis | 0.1 (1) | 0 | 0 |
| Comorbidities | Hypertension | 22.3 (187) | 50 (2) | 40 (10) |
|  | T2DM | 4.3 (36) | 0 | 12 (3) |
|  | Hypothyroidism | 11.4 (96) | 0 | 20 (4) |
|  | Asthma | 1.1 (9) | 0 | 0 |
|  | Osteoporosis | 2.9 (24) | 0 | 0 |
|  | CKD | 0.1 (1) | 0 | 0 |
|  | D(I)CM | 1.1 (9) | 25 (1) | 0 |
|  | CLD | 0.1 (1) | 0 | 0 |
| Drugs | Prednisolone | 12.14 (102) | 0 | 28 (7)<sup>a</sup> |
|  | Hydroxychloroquine | 33 (278) | 75 (3) | 40 (10) |
|  | Methotrexate | 60.4 (507) | 75 (3) | 68 (17) |
|  | Sulfasalazine | 17.26 (145) | 0 | 12 (3) |
|  | Leflunomide | 24 (203) | 25 (1) | 44 (11) |
|  | MMF | 3.7 (31) | 0 | 4 (1) |
|  | Azathioprine | 3.9 (33) | 0 | 4 (1) |
|  | Etanercept | 0.8 (7) | 0 | 0 |
|  | Infliximab | 1 (8) | 0 | 0 |
|  | Adalimumab | 0.5 (4) | 0 | 4 (1)<sup>b</sup> |
|  | Golimumab | 1.2 (10) | 0 | 4 (1)<sup>c</sup> |
|  | Rituximab | 1.3 (11) | 0 | 4 (1) |
|  | Tocilizumab | 0.2 (2) | 0 | 0 |
|  | Tofacitinib | 0.6 (5) | 0 | 0 |
|  | Filgotinib | 0.5 (4) | 0 | 0 |

Numbers within individual cells represent either percentage (number) or mean ± standard deviation
a Prednisolone use in asymptomatic 11.71% (95/811) vs 28% (7/25) in patients with flu-like symptoms, $p=0.024$
b Adalimumab use in asymptomatic 0.37% (3/811) vs 4% (1/25) in patients with flu-like symptoms, $p=0.019$
c Golimumab use in asymptomatic 1.11% (9/811) vs 4% (1/25) in patients with flu-like symptoms, $p=0.048$
All of these p-values are above cut-off set after correcting for family wise error rate, i.e., 0.0166, and therefore non-significant
All other bivariate comparisons have $p\text{-values} > 0.05$
Abbreviation: AS: ankylosing spondylitis; CKD: chronic kidney disease; CLD: chronic liver disease; MMF: mycophenolate mofetil;
PsA: psoriatic arthritis; RA: rheumatoid arthritis; SLE: systemic lupus erythematosus; SjS: Sjogren syndrome; SSc: scleroderma;
T2DM: type 2 diabetes mellitus;

### COVID-19 cases

Among our patients 29 reported flu-like symptoms but rT-PCR on nasophayngeal swab was done only on six cases (four with RA, one with AS, and one with SLE) among which four tested positive and the rest didn’t report to the medical facility. Detailed demographic and clinical features of COVID-19 positive cases is given in Table 2. None of the patients were on tsDMARDs or monoclonal antibodies. Two had hypertension and one had coronary artery disease as comorbidities. All of them were admitted in Government run medical institutions with fever. None of the patients required intensive care or ventilatory support. All were discharged without any complications. All of these patients temporarily discontinued their rheumatological drugs, except hydroxychloroquine. After recovery from COVID-19, one patient with RA had a disease flare which resolved with institution of DMARDs.

**Table 2.**
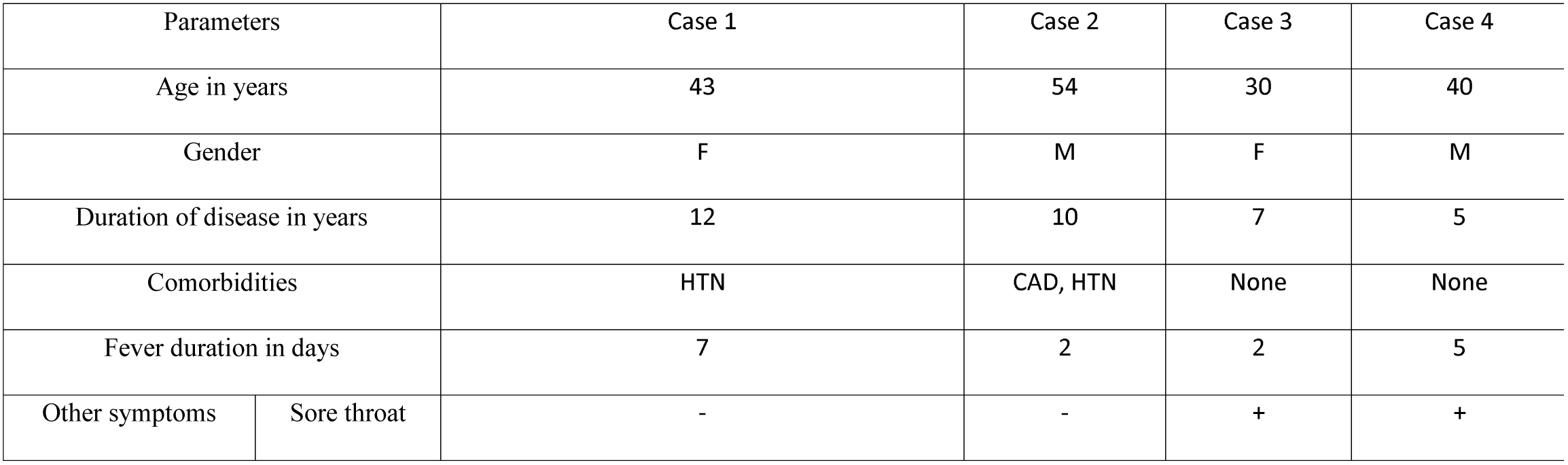
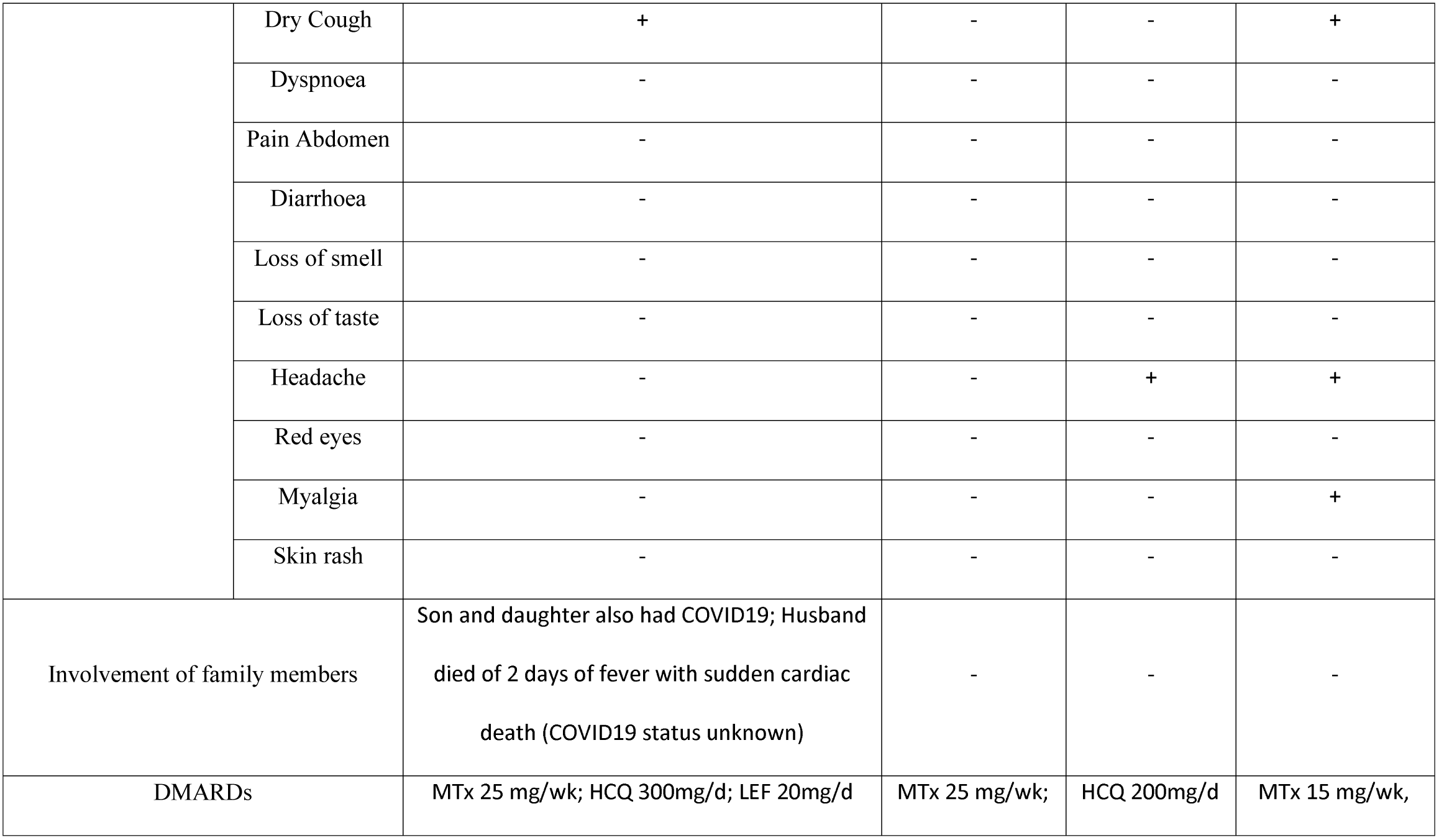
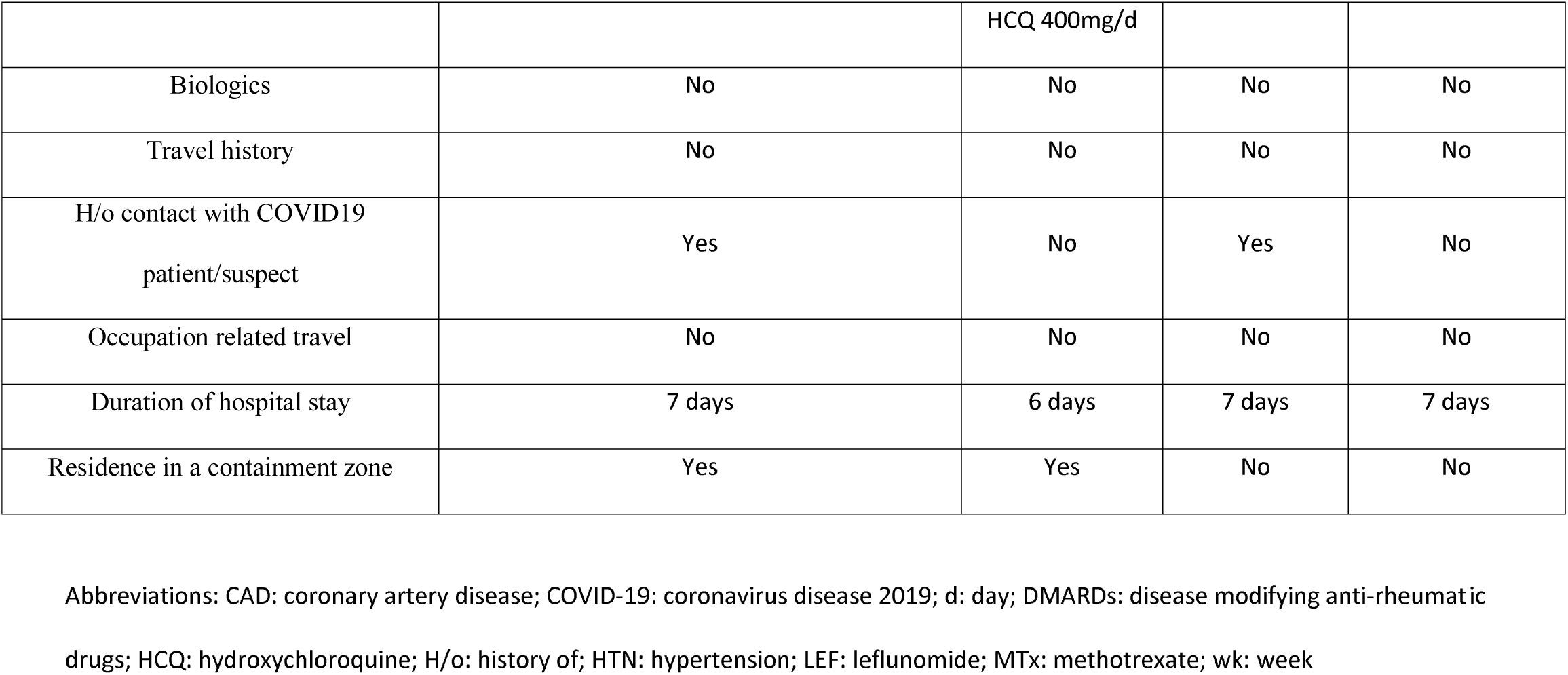
Details of individual cases of definite COVID-19 patients

The incidence of confirmed COVID-19 in our cohort was 0.476% (95% CI 0.19-1.22). The background general population living in Delhi during this period is 21777954, [**7**] and number of COVID-19 cases in Delhi during the study period was 113193, [**8**] giving incidence of 0.519% (95% CI: 0.517-0.523). The incidence of COVID-19 was similar to general Delhi population (0.476% vs 0.519%, difference 0.043%, 95% CI: −0.69 to 0.33, p=0.86). Incidence of COVID-19 cases was asymmetrically distributed among different diagnostic classes (Figure 1). If we consider only the diagnostic class of RA, even then the incidence of COVID-19 was similar between patients with RA and the general Delhi population (0.56% vs 0.519%, difference 0.041%, 95% CI: −0.3012% to 0.9131%, p=0.87).

### COVID-19: clinically suspected cases

Another 25 patients, had flu-like symptoms but were not tested with nasopharyngeal swab. None of these patients reported travel outside India or Delhi or even contact with a known COVID-19 case. Details of these patients are summarized in Table 1. However, all of them resided within containment zones, which are zones demarcated by the Government as high risk of transmission and have strong restriction of movement. Twenty-one of them had RA, three had SLE and one had AS. Among these patients, seventeen were on methotrexate, eleven were on leflunomide, ten were on hydroxychloroquine, seven were on prednisolone, three on sulfasalazine, two on anti-TNF agents and one each on azathioprine, mycophenolate and rituximab. Only one of these patients was hospitalized and needed supplemental oxygen but made a full recovery. The incidence of Flu-like symptoms in patients with RA was similar to those observed in CTD (2.95%, 21/713 vs 2.8%, 3/107 respectively, p=0.999) and patients with SpA (2.95%, 21/713 vs 5%, 1/20, respectively, p=0.46).

Two patients with RA had stopped medications and both of them and four other patients with RA experienced a disease relapse, all of them after the flu-like episode. One patient with SLE also discontinued all her medications following her flu-like episode and developed a renal flare with subnephrotic proteinuria and raised creatinine for which she was instituted on high dose prednisolone and mycophenolate and is still in re-induction therapy.

## Discussion

This is the first detailed report in a cohort of patients with various rheumatological diagnoses from an area of high transmission and case load from India. We observed a similar incidence of COVID-19 among patients with rheumatological diseases compared to the general population of Delhi. The results were stable even after the sensitivity analyses done to reduce heterogeneity. However, between patients COVID-19 positive and negative cases, the distribution of immunosuppressive drugs was similar as were the comorbidities. All the COVID-19 infections were uncomplicated. Increased risk of infection, especially respiratory viral infections, occur with increased frequency among patients with rheumatic diagnoses especially those on immunosuppressive therapy [9, 10]. However, two previous studies from Italy, in the same tune as ours, comparing incidence of COVID-19 infection on biologics or tsDMARDs with general population, found no significant differences [**1, 11**]. The range of reported incidence among patients with rheumatic diseases was 0.38-0.62% which is similar to what we have observed. All of our patients had RA. On the other hand, a previous Indian report cited one case of COVID-19 among 845 patients with lupus [12]. However, among this cohort 17 had suspect symptoms, among which only two were tested. Therefore, the incidence of flu-like symptoms in that cohort was 2%, which is very similar to what was observed among our patients with CTD (2.8%).

Regarding the impact of COVID-19 on our patients, we observed unscheduled stoppage of medications in 15.7% (132/840) patients and increased disease activity in 2% (16/840) of patients. One quarter of our patients relapsed after recovery from COVID-19 infection and 28% (7/25) of our other patients with flu-like symptoms relapsed after recovery. Total post infectious rheumatic disease relapse rate was 27.6%, 95% confidence interval 14.7-45.7). A myriad of mechanisms is hypothesized for this kind of reactivation of autoimmune disease after COVID-19 ranging from molecular mimicry, bystander killing, epitope spreading, viral persistence to formation of neutrophil extracellular traps [13]. However, the most plausible explanation is stoppage of immunosuppressive drugs during the infectious episode. Recent evidence suggests that dexamethasone in COVID-19 is actually useful for those on mechanical ventilation or oxygen support and at least not harmful for others not on oxygen support [14]. Therefore, it might not be a bad measure, not to stop the glucocorticoids during COVID-19 infection especially among patients with RA or SLE.

Our study has a few limitations: single-centre cross-sectional survey, less than a 100% response to the telephonic survey and all suspected patients were not confirmed with rt-PCR. However, long-term follow up and detailed diagnostic and therapeutic information and details of flu-like symptoms and COVID-19 natural history over repeated and multiple telephonic conversations with the patients are the strengths of our study. To conclude, we observed that the incidence of COVID-19 infection among patients with rheumatic diseases was similar to the community. The incidences of Flu-like symptoms were similar across different diagnostic classes.

DMARDs neither increased risk of the infection nor provided any protection to the same. Since, relapse of disease, especially after stoppage of disease modifying agents, is a concern after this infection, continuation of glucocorticoid through the infection might be a prudent strategy.

## Data Availability

Data is available with the corresponding author

## Compliance with ethical standards

The study protocol was reviewed and approved by the institutional review board (no. IEC-280/17.4.2020).

## Funding

Funded by Institute Research Project, All India Institute of Medical Sciences, New Delhi, **No.F.5-59/IRG/2010/RS**

## Declarations

Conflicts of interest/Competing interests:

none to declare

### Consent to participate

Informed consent taken from each participant

### Availability of data and material

Data is available with the corresponding author and is freely available on request

## Authors’ contributions

RPG: conception, data acquisition and analysis, drafting, revising, final approval of the manuscript UK: conception, revising, final approval

MK: data acquisition, revising, final approval

DR: data acquisition. revising, final approval

PV: data acquisition. revising, final approval

DKM: data acquisition. revising, final approval

LM: data acquisition. revising, final approval

SN: data acquisition. revising, final approval

DB: data acquisition. revising, final approval All authors agree to be accountable for all aspects of the work including accuracy of the data and integrity

## Notes

### Competing Interest Statement

The authors have declared no competing interest.

### Author Declarations

The study protocol was reviewed and approved by the institutional review board of AIIMS, New Delhi (no. IEC-280/17.4.2020).

